# Diagnostic Delays Drive Transmission in Dense Cities: Modeling the Waiting-Window Effect and Its Mitigation

**DOI:** 10.64898/2026.04.20.26350946

**Authors:** S. Bahig, M. Oughton, J. Vandesompele, I. Brukner

## Abstract

In dense urban settings, delays between diagnostic sampling and effective isolation can sustain transmission during peak infectiousness. We define a waiting-window transmission externality arising when infectious individuals remain mobile while awaiting results, formalized as E = N·P·TR·D, where N is daily testing volume, P test positivity, TR transmission during the waiting period, and D turnaround time. Using Monte Carlo simulation and a susceptible-infectious-recovered (SIR) framework, we quantify excess infections per 1,000 tests/day under multiple diagnostic workflows. A surge scenario incorporates positive coupling between TR and D (ρ = 0.45), reflecting co-occurrence of laboratory saturation and elevated contacts during system stress. Under centralized 48-hour workflows, excess infections reach ∼80 at P = 10% and ∼401 at P = 50%, increasing to ∼628 under surge conditions. In contrast, near-patient rapid testing and home sampling reduce this to ∼5 and ∼25–26, respectively. Workflows that eliminate the waiting window—either through immediate isolation at sampling or through home-based PCR that returns results at the point of collection—effectively collapse the transmission term. These findings identify diagnostic delay as a modifiable driver of epidemic dynamics. Operational redesign of testing workflows, including decentralized sampling and home-based molecular diagnostics, offers a scalable pathway to improve epidemic controllability and reduce inequities in dense urban environments.

## 1. Introduction

Epidemic control in dense urban environments depends critically on the timing of intervention relative to infectiousness. For respiratory pathogens such as SARS-CoV-2, peak transmissibility occurs near or before symptom onset, rendering delays between diagnostic sampling and effective isolation epidemiologically consequential. While testing is typically framed as a detection tool, its operational characteristics—particularly turnaround time and associated behavior during result waiting—can themselves shape transmission dynamics. This creates a paradox: diagnostic systems intended to mitigate spread may inadvertently sustain it when delays are present, especially under high-incidence conditions.

This work examines the impact of operational changes in sampling strategies and quarantine protocols on C_case, defined as the break-even cost per infection averted. By modifying these processes, such adjustments directly influence the economic efficiency of surveillance and control workflows, enabling more informed cost-effectiveness evaluations during periods of high transmission pressure (e.g., epidemic surges).

Centralized diagnostic systems frequently introduce operational delay and non-contact-free logistics (travel, transit, and indoor queues), particularly during surges when testing demand, positivity, and crowding increase. In this setting, public health guidance may recommend precautionary quarantine, but effective isolation is often delayed until results are returned, creating a multi-day ‘waiting window’ during which mobile infectious individuals can continue transmitting. This creates a specific failure mode for dense cities: the control system becomes least effective exactly when incidence rises. This paper quantifies how correlated uncertainty in transmission rate (TR) and turnaround time (D) during surges amplifies risk, and demonstrates that home sampling remains cost-effective even at higher per-test prices.

The WHO CORC-CoV (Coronavirus Collaborative Open Research Consortium) roadmap highlights the limited availability of rapid, multiplex point-of-care platforms and calls for low-cost, high-performance diagnostics to enable earlier public health action (World Health Organization, 2026). Here, we formalize a simple ‘waiting-window’ transmission externality induced by test turnaround time, quantify its scaling with test positivity and delay, and outline an operational pathway for home sampling that triggers precautionary isolation at sampling, to collapse this externality while maintaining result interpretability through sample adequacy control (SAC).

## 2. Methods

### 2.1. Waiting-window transmission externality

Let N denote the number of tests performed per day (tests/day), P the test positivity (fraction of tests that are positive; proxy for the fraction of tested individuals who are infectious), TR the expected transmissions per infectious person per day during the waiting period (residual contacts while awaiting results), and D the turnaround time from sampling to an actionable result (days). The waiting-window contribution to transmission is proportional to P·TR·D.

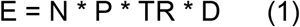

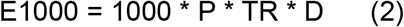

Because epidemics also include background transmission not prevented by the testing workflow, we represent that component with an additive baseline term X. Fig. 1 displays the y-axis as X + E_1000_, with tick labels written as X, X+200, X+400, etc.

**Fig. 1.**
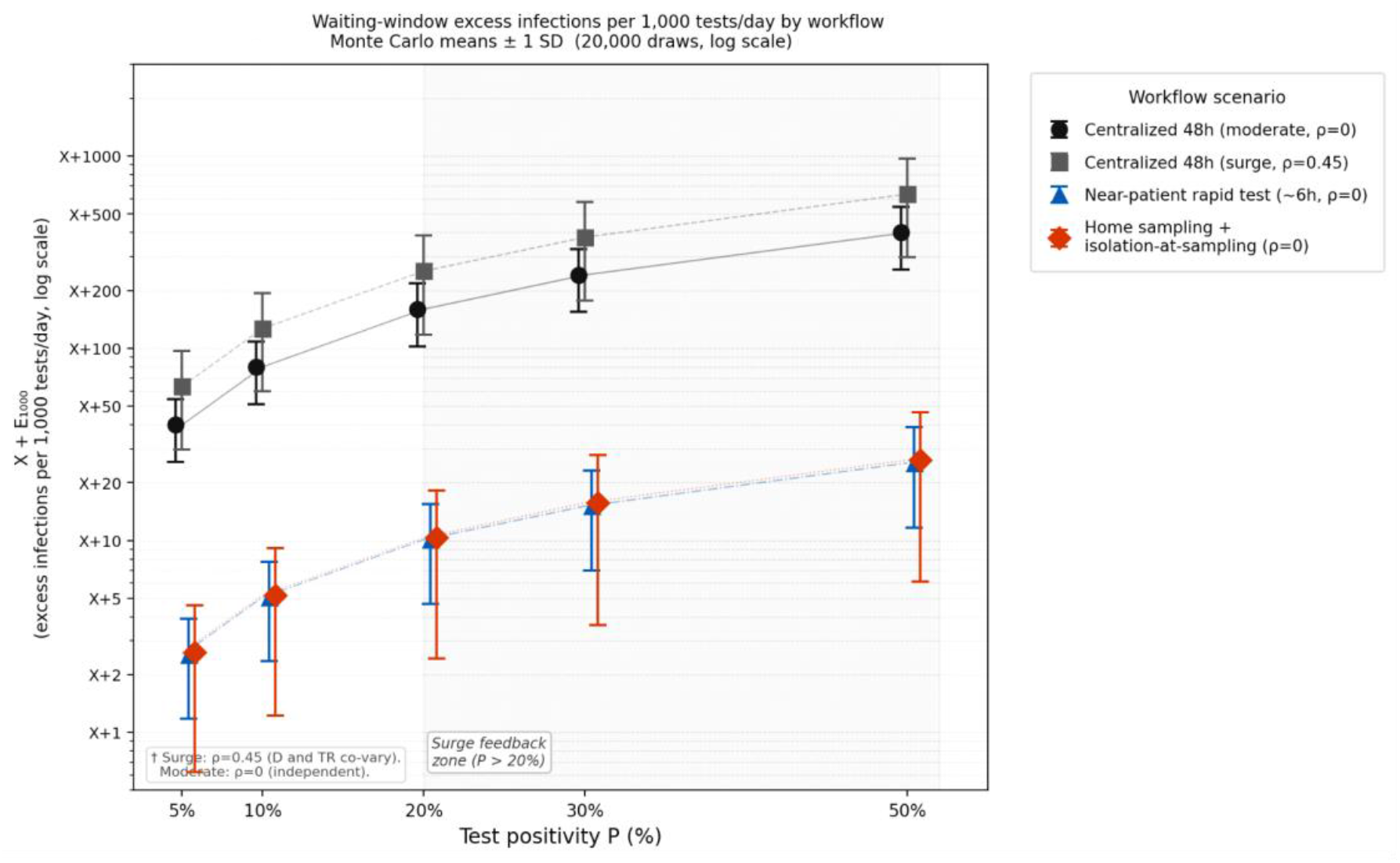
Waiting-window excess infections per 1,000 tests/day by workflow (logarithmic scale). Discrete data points represent Monte Carlo means (20,000 iterations); error bars = ±1 SD. Logarithmic y-axis ensures near-patient and home-sampling scenarios (near X) are clearly distinguishable from centralized scenarios. Circles: centralized 48h moderate (ρ=0); Squares: centralized 48h surge (ρ=0.45, Gaussian copula, wider bars reflect co-occurrence of lab saturation D↑ and crowding risk TR↑); Triangles: near-patient rapid test (∼6h, ρ=0); Diamonds: home sampling + isolation-at-sampling (ρ=0). Legend positioned outside plot area. Shaded region (P>20%):H surge feedback zone, characterized by sudden, massive influx of patients that exceeds normal operating capacity. Y-axis displayed as X+E_1000_ to avoid implying zero background incidence.

**Fig. 2.**
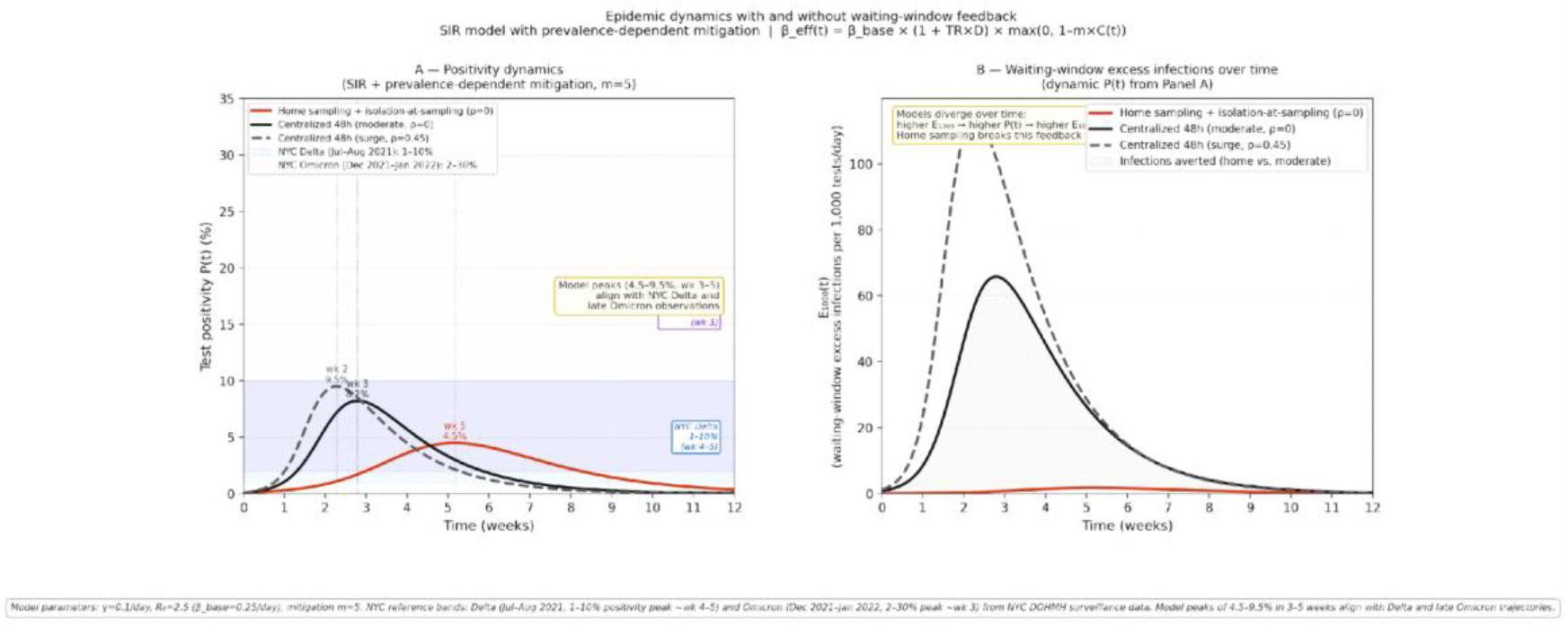
Epidemic dynamics with and without waiting-window feedback (SIR model with prevalence-dependent mitigation, β_eff(t) = β_base × (1 + TR×D) × max (0, 1–m×C(t))). Panel A: test positivity P(t) over 12 weeks, with NYC real-world reference bands overlaid: Delta wave (Jul–Aug 2021, 1–10% peak, ∼wk 4–5, blue band) and Omicron wave (Dec 2021–Jan 2022, 2–30% peak, ∼wk 3, purple band). Model peaks of 4.5–9.5% in 3–5 weeks align with Delta and late Omicron trajectories. Panel B: E_1000_(t) over 12 weeks. Parameters: γ=0.1/day, R_0_=2.5, m=5. The qualitative divergence between centralized and home-sampling workflows is robust across this parameter range.

### 2.2. Parameter evidence and operational measurement

P can be operationalized using test positivity, a standard surveillance indicator routinely reported by public health systems (CDC, 2025). D is the operational sample-to-action turnaround time; observational analyses of COVID-19 RT-PCR workflows have shown that delays can be socially patterned (Núñez et al., 2022). Although a pathogen’s intrinsic infectivity rate (e.g., R_0_) governs the baseline speed of spread, TR is a distinct and operationally modifiable parameter; unlike R_0_, which reflects pathogen biology, TR is determined by human behavior, mobility decisions, queuing, and contact patterns during the waiting period. TR captures residual mobility and contact behavior while awaiting results, including travel and queue exposures when testing is centralized, and is therefore amenable to intervention through workflow design rather than pathogen biology. Early infectiousness around symptom onset implies that even short delays can preserve a substantial fraction of transmissibility (He et al., 2020). Evidence syntheses indicate that a substantial fraction of transmission arises from people without symptoms (including those infected before being symptomatic), motivating inclusion of a background term X (Oran and Topol, 2020; Buitrago-Garcia et al., 2020; Johansson et al., 2021).

### 2.3. Parameter variability and uncertainty propagation

We represent TR and D using truncated normal distributions (lower bound 0) and propagate uncertainty by Monte Carlo sampling (20,000 draws). To better reflect real-world surge dynamics, we introduced a positive correlation coefficient (ρ=0.45) for the surge scenario, indicating that laboratory saturation (increased D) typically coincides with increased waiting-period contacts (increased TR) due to system-wide pressure, longer queues, crowding at testing sites, and delayed isolation all co-occur. The moderate scenario retains ρ=0 because, under normal operational conditions, laboratory throughput and individual contact behavior vary independently; only when the system saturates do these parameters become coupled. The copula is implemented via Cholesky decomposition of the 2×2 correlation matrix, with marginals mapped through the truncated normal inverse CDF.

### Scenario parameterizations (mean ± SD)

- Centralized 48h (moderate, ρ=0): TR=0.40±0.10/day; D=2.0±0.5 days.
- Centralized 48h (surge, ρ=0.45): TR=0.60±0.15/day; D=2.0±0.8 days; ρ=0.45 via Gaussian copula.
- Near-patient rapid test (∼6h, ρ=0): TR=0.20±0.07/day; D=0.25±0.10 days.
- Home sampling + isolation-at-sampling (ρ=0): TR=0.02±0.02/day; D=2.0±0.8 days.

These values are illustrative, spanning plausible operational states in dense cities. Fig. 1 uses a logarithmic y-axis and discrete data points with ±1 SD error bars. Because E is a non-negative product of random variables, Monte Carlo uncertainty intervals can be mildly right-skewed, as visible in the asymmetric SD bars at high P in Fig. 1.

### 2.4. Illustrative break-even cost per infection

The break-even cost per infection (C_case*) is calculated as:

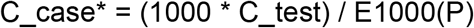

#### Where

- C_test = $40 is the cost per test
- E1000(P) is the expected number of excess infections per 1,000 tests at prevalence P

#### Interpretation

- When the societal cost per infection exceeds C_case*, a testing workflow that removes the waiting-window term is economically favorable.
- C_case* is a threshold, not a direct estimate of cost.

#### Examples

- At P = 10%: C_case* = $500
- At P = 50% (surge conditions): C_case* = $100

Applying standard severity weights (∼2% hospitalization, ∼0.5% fatality), this corresponds to approximately $26,300 per hospitalization prevented and $105,000 per death averted at P = 10%, which is within commonly cited cost-effectiveness thresholds ($50,000–$150,000 per Quality-Adjusted Life Year (QALY)) used by the National Institute for Health and Care Excellence (NICE, UK) (National Institute for Health and Care Excellence, 2013) and the Canadian Agency for Drugs and Technologies in Health (CADTH, Canada) (Canadian Agency for Drugs and Technologies in Health, n.d.).

## 3. Results

### 3.1. Positivity–delay scaling and scenario comparison

Fig. 1 shows X + E_1000_ as a function of test positivity P on a logarithmic scale, displayed as discrete data points with ±1 SD error bars (20,000 Monte Carlo draws). The surge scenario (ρ=0.45) shows markedly wider error bars than the moderate scenario (ρ=0), reflecting dangerous unpredictability when lab saturation and crowding co-occur. A surge feedback zone (P>20%) is annotated.

### 3.2. Monte Carlo estimates for representative workflows

Table 1 summarizes daily E_1000_ across representative positivity values (P) for all four workflow scenarios. Under the moderate 48h workflow (ρ=0), E_1000_ is ∼80±29 at P=10% and ∼401±144 at P=50%. Under surge mixing with TR-D correlation (ρ=0.45), E_1000_ rises to ∼126±67 at P=10% and ∼628±334 at P=50% — the substantially wider SD is the critical signal, reflecting unpredictability during system saturation. The near-patient rapid test yields ∼5±3 at P=10% and ∼25±14 at P=50%, epidemiologically equivalent to home sampling (∼5±4 and ∼26±20), confirming that either reducing D or reducing TR effectively collapses the waiting-window term.

**Table 1.**
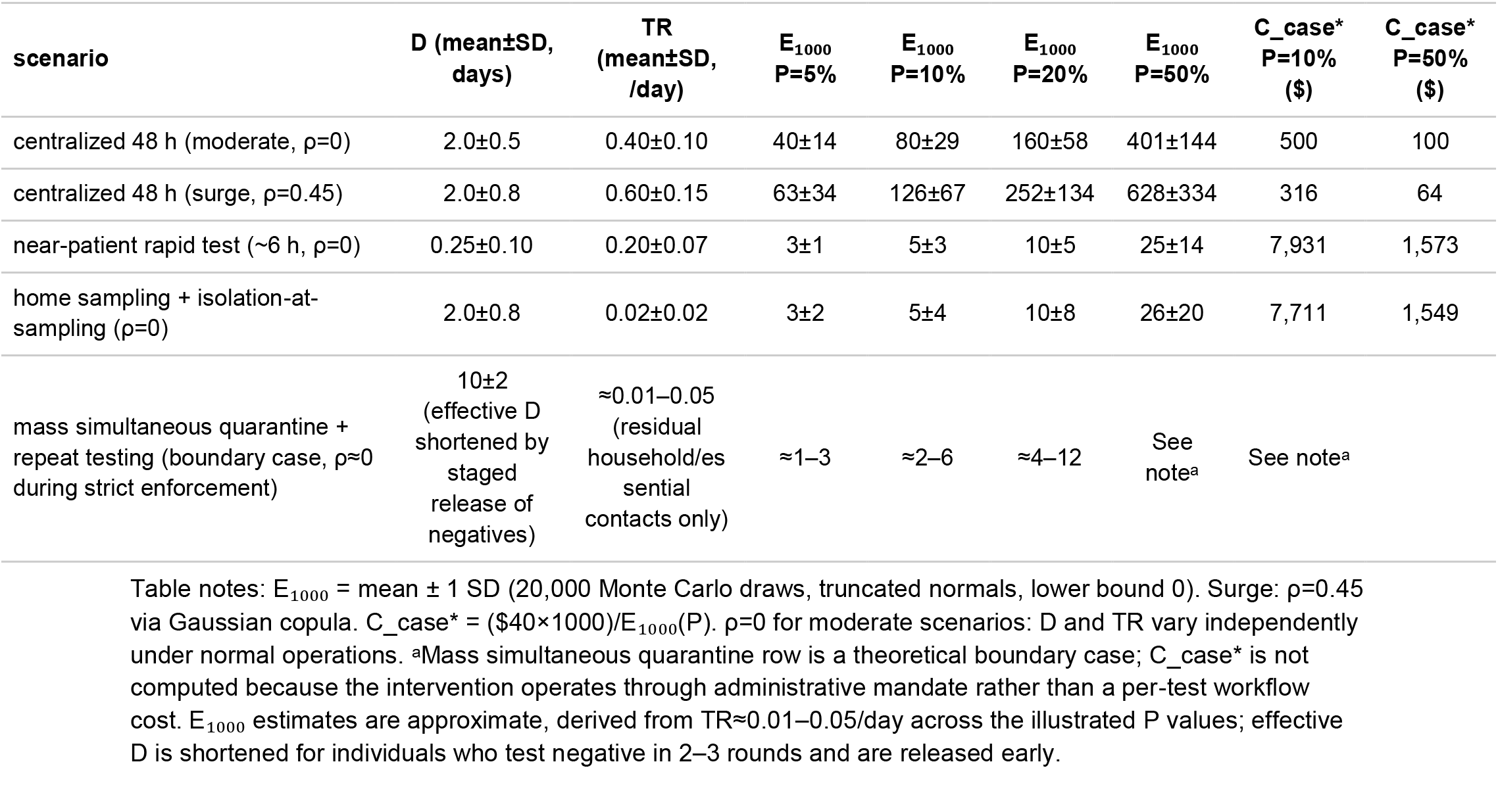
Waiting-window excess infections and C_case* per 1,000 tests/day under representative positivity (P)

## 4. Discussion

### 4.1. Epidemiological and operational implications

This study identifies diagnostic delay as a previously under-formalized driver of transmission in dense urban settings. The waiting-window externality emerges not from pathogen biology but from the interaction between testing workflows and human behavior during the result interval. Critically, this effect scales with test positivity and becomes amplified under surge conditions, when laboratory delays and contact patterns become coupled. As a result, centralized diagnostic systems may become least effective precisely when epidemic control is most needed.

When home sampling is combined with precautionary isolation at the time of collection, TR is reduced toward zero during the waiting window, preventing the delay-induced transmission externality and improving epidemic controllability.

A useful boundary comparator is mass simultaneous quarantine, the population-wide extreme of the same logic underlying home sampling. When an entire population is mandated to stay home during a defined intervention window (as implemented during early COVID-19 containment efforts in China and several other jurisdictions), TR is driven toward near zero across the entire population (TR ≈ 0.01–0.05/day for residual household and essential-worker contacts), even more aggressively than home sampling (TR = 0.02±0.02/day in our parameterization). In the SIR framework, this can be represented as a short, time-limited “hard reset”: β^eoo^(t) → ≈0 for approximately 10 days, after which transmission resumes but starts from a substantially lower infectious pool, reflecting clearance achieved through 2–3 repeated testing rounds (with negatives released early, reducing effective D per individual). This boundary case thus illustrates the upper bound of TR suppression: it confirms that collapsing TR is the operative epidemiologic mechanism, while simultaneously demonstrating why home sampling is the scalable and less disruptive pathway to the same effect — achieving TR reduction without the severe socioeconomic costs, logistical demands, and limitations on individual liberty associated with city-wide coercive quarantine.

The SIR model predictions are consistent with real-world observations: during the NYC Delta wave (July–August 2021), test positivity peaked at 5–10% over approximately 4–5 weeks; during the NYC Omicron wave (December 2021–January 2022), positivity rose from ∼2–3% to ∼30% over ∼3 weeks, doubling every 5–7 days before behavioral change and policy responses dampened growth. Our model produces peaks of 4.5–9.5% in 3–5 weeks under the calibrated mitigation parameter (m=5), aligning with the Delta and late Omicron trajectories. This consistency supports the plausibility of the modelled workflows, while the qualitative divergence between centralized and home-sampling scenarios remains robust across this parameter range (NYC DOHMH, 2022).

### 4.2. Economic thresholds and outcome-based analysis

C_case* is a break-even threshold, not a cost estimate. The transition from case-based metrics to outcome-based economics allows for direct benchmarking against international health standards. Centralized testing C_case* collapses during surges (from $500 to $100 at P=50%), reflecting operational fragility. Home sampling C_case* remains high, a sign of success, as it means the workflow can cost considerably more per test and still be economically rational.

Beyond C_case*, applying standard severity weights (∼2% hospitalization, ∼0.5% fatality) indicates approximately $26,300 per hospitalization prevented and $105,000 per death averted at P=10% well within the $50,000–$150,000/QALY thresholds used by National Institute for Health and Care Excellence (2013) and Canadian Agency for Drugs and Technologies in Health (n.d.). During 2020–2022, governments implicitly valued preventing one hospitalization at tens to hundreds of thousands of dollars, far above these thresholds.

### 4.3. Socioeconomic dynamics of quarantine breakdown

In high-density settings with precarious work, limited paid sick leave, and crowded housing, prolonged waiting windows can make quarantine non-viable, driving non-adherence, delayed testing, and hidden transmission. At scale, this triggers a feedback loop: higher incidence → higher positivity and longer turnaround → higher E_1000_ → further incidence growth. From a governance perspective, sustained infeasible quarantine can erode trust or trigger escalation toward coercive enforcement, both carrying substantial social and economic costs. Home sampling paired with SAC and test-triggered precautionary isolation reduces the need for coercion by shortening the operational waiting window.

Behavioral evidence from COVID-19 indicates that adherence to test-trace-isolate recommendations is often constrained by feasibility and burden (Smith et al., 2021). Diagnostic delay is frequently driven primarily by test turnaround time and is socially patterned, with longer delays associated with socioeconomic marginalization (Núñez et al., 2022). Delay is not a neutral operational detail; it is an equity-sensitive amplification factor during surges.

### 4.4. Feasibility and implementation pathway

Home sampling does not require a single centralized laboratory bottleneck. In dense cities, a scalable model is a hub-and-spoke workflow: (i) self-collection at home using SAC to make negative results interpretable (Brukner et al., 2020, 2021); (ii) point-of-collection use of validated inactivating/stabilizing transport buffers has been evaluated for RNA stability and compatibility with extraction-free RT-qPCR and rapid antigen workflows (Claeys et al., 2024; Brukner and Oughton, 2025); (iii) decentralized local molecular testing running RT-qPCR on standard instruments with multiple daily runs; and (iv) sample pooling to keep costs and turnaround times maximally under control. Under this design, sample-to-action times of ∼1– 12 hours are operationally plausible. (Verwilt et al., 2022).

Stabilized, inactivated specimens enable practical collection logistics, including once- or twice-daily front-door pickup or neighborhood drop boxes (Ciccone et al., 2021). Pre-distributed home sampling kits, when paired with barcode/QR-based reporting and support services (paid sick leave, isolation support), improve feasibility, reduce reliance on coercive enforcement, and stabilize both epidemic control and socioeconomic continuity. For completeness, home-based PCR platforms—defined as self-contained molecular diagnostic systems that deliver results at the point of sampling—represent an epidemiologically equivalent strategy to precautionary isolation at sampling. By collapsing turnaround time to minutes, these systems eliminate the waiting window while simultaneously removing uncertainty around infection status. From a transmission perspective, this is equivalent to driving waiting-period transmission toward zero because individuals can act immediately on confirmed results. Although current costs and scalability remain limiting, home-based PCR reinforces the central operational principle demonstrated in this study: minimizing turnaround time and waiting-period transmission is the most effective strategy for reducing epidemic spread.

## 5. Limitations

This analysis uses a deliberately simple multiplicative model to isolate the effect of operational delay on transmission. Test positivity P is used as a proxy for the fraction infectious among those tested and will vary with testing strategy and case ascertainment. The surge scenario explicitly models positive TR-D correlation (ρ=0.45) via a Gaussian copula; this value is illustrative and sensitivity analyses with alternative values of ρ are straightforward using the provided simulation code. The moderate scenario deliberately retains ρ=0 to reflect normal operational independence. C_case* uses a fixed $40/test and ignores broader benefits (equity, adherence) and real-world cost variation ($60–$80/test for home sampling). Secondary transmission, heterogeneity in contact networks, and behavioral feedback are not explicitly modelled. The baseline term X is conceptual and not estimated. Despite these simplifications, the linear dependence of E_1000_ on P and D highlights a robust operational mechanism: delay creates a transmission externality that worsens exactly when systems are stressed. Three additional boundary conditions warrant explicit acknowledgement. First, at very low positivity (P < 5%), E_1000_becomes insensitive to TR and D: the waiting-window term is small regardless of workflow, and centralized testing is operationally adequate at this threshold. The model therefore does not claim that home sampling and temporary isolation is universally superior, only that it becomes critical above a certain positivity level. Second, if TR cannot realistically approach zero due to within-household transmission (floor ∼0.05– 0.10/day), the home sampling advantage shrinks but holds: household transmission is structurally different from queue or transit exposure, it is bounded, more predictable, and not amplified by centralized logistics. Third, at high P, TR and P may not be fully independent, elevated positivity could alter individual contact behavior in ways not captured by the current parameterization, representing a direction for future empirical work. Finally, while mass simultaneous quarantine is included as a theoretical boundary case showing the upper bound of TR suppression, its real-world implementation introduces correlations not modelled here: testing backlogs, enforcement heterogeneity, and essential-worker exemptions could elevate effective TR above the theoretical floor, and the severe socioeconomic disruption of city-wide coercive quarantine lies outside the scope of this analysis.

## 6. Conclusions

In dense cities, centralized testing with multi-day turnaround creates a delay-driven, self-amplifying transmission term that scales with positivity and correlated TR-D uncertainty. Home sampling coupled with precautionary isolation at sampling can collapse this term and reduce the likelihood of quarantine breakdown during surges, while sample adequacy control, validated inactivation/stabilization buffers, and home-based PCR support scalable low-delay workflows. The C_case* metric demonstrates that investing in low-delay diagnostic strategies is not only epidemiologically sound but also cost-effective when systems are most stressed. These operational design choices offer a practical pathway to maintain epidemic controllability and mitigate inequities during future respiratory-pandemic surges.

## Consent for publication

Not applicable.

## Data availability

All simulation parameters and scenario definitions are provided in the manuscript. Python simulation code is available from the corresponding author on reasonable request.

## Author contributions (CRediT)

Conceptualization: IB, MO. Methodology: IB, SB. Formal analysis: IB, SB. Visualization: IB, SB. Writing - original draft: SB, IB. Writing - review and editing: SB, IB, MO, JV.

## Declaration of generative AI and AI-assisted technologies in the manuscript preparation process

During the preparation of this work the authors used ChatGPT and Claude to assist with language editing, notation consistency, and simulation code development. After using these tools, the authors reviewed and edited the content as needed and take full responsibility for the content of the published article.

## Acknowledgements

We thank Benoit Houle, Montreal, for advice and encouragement to visualize different pandemic scenarios. Microbiology Department members at JGH, Montreal, shaped pandemic discussions in the past, affecting this work.

## References

Brukner, I., Eintracht, S., Papadakis, A.I., et al., 2020. Maximizing confidence in a negative result: Quantitative sample adequacy control. J. Infect. Public Health 13 (7), 991–993.

Brukner, I., Oughton, M., 2025. Direct PCR for Rapid and Safe Pathogen Detection: Laboratory Evaluation Supporting Field Use in Infectious Disease Outbreak. LabMed 2 (3), 12. 10.3390/labmed2030012.

Brukner, I., Resendes, A., Eintracht, S., et al., 2021. Sample Adequacy Control (SAC) lowers false negatives and increases the quality of screening. Diagnostics 11 (7), 1133.

Buitrago-Garcia, D., Egli-Gany, D., Counotte, M.J., et al., 2020. Occurrence and transmission potential of asymptomatic and presymptomatic SARS-CoV-2 infections. PLOS Med. 17 (9), e1003346.

Canadian Agency for Drugs and Technologies in Health, n.d. Guidelines for Economic Evaluation of Health Technologies.

Centers for Disease Control and Prevention, 2023. CDC 2019-Novel Coronavirus Real-Time RT-PCR Diagnostic Panel.

Centers for Disease Control and Prevention, 2025. COVID-19 Surveillance Data-Test Positivity.

Ciccone, E.J., Conserve, D.F., Dave, G., et al., 2021. At-home testing to mitigate community transmission of SARS-CoV-2. BMC Public Health 21, 2209.

Claeys, M., Al Obaidi, S., Bruyland, K., et al., 2024. Assessment of DNA/RNA Defend Pro. Int. J. Mol. Sci. 25 (16), 9097.

Cutler, D.M., Summers, L.H., 2020. The COVID-19 pandemic and the $16 trillion virus. JAMA 324 (15), 1495–1496.

He, X., Lau, E.H.Y., Wu, P., et al., 2020. Temporal dynamics in viral shedding and transmissibility of COVID-19. Nat. Med. 26, 672–675.

Hellewell, J., Abbott, S., Gimma, A., et al., 2020. Feasibility of controlling COVID-19 outbreaks by isolation of cases and contacts. Lancet Glob. Health 8 (4), e488–e496.

Johansson, M.A., Quandelacy, T.M., Kada, S., et al., 2021. SARS-CoV-2 transmission from people without COVID-19 symptoms. JAMA Netw. Open 4 (1), e2035057.

Kretzschmar, M.E., Rozhnova, G., Bootsma, M.C.J., et al., 2020. Impact of delays on effectiveness of contact tracing strategies for COVID-19. Lancet Public Health 5 (8), e452–e459.

Larremore, D.B., Wilder, B., Lester, E., et al., 2021. Test sensitivity is secondary to frequency and turnaround time for COVID-19 screening. Sci. Adv. 7 (1), eabd5393.

Mina, M.J., Parker, R., Larremore, D.B., 2020. Rethinking COVID-19 test sensitivity-a strategy for containment. N. Engl. J. Med. 383 (22), e120.

Mullachery, P.H., Li, R., Melly, S., et al., 2022. Inequities in spatial accessibility to COVID-19 testing in 30 large U.S. cities. Soc. Sci. Med. 310, 115307.

National Institute for Health and Care Excellence, 2013. Guide to the methods of technology appraisal.

Nunez, I., Belaunzaran-Zamudio, P.F., Caro-Vega, Y., 2022. Result turnaround time of RT-PCR for SARS-CoV-2. Rev. Invest. Clin. 74 (2), 71–80.

Oran, D.P., Topol, E.J., 2020. Prevalence of asymptomatic SARS-CoV-2 infection. Ann. Intern. Med. 173 (5), 362–367.

Rader, B., Astley, C.M., Sy, K.T.L., et al., 2020. Geographic access to United States SARS-CoV-2 testing sites highlights healthcare disparities. J. Travel Med. 27 (7), taaa076.

Smith, L.E., Potts, H.W.W., Amlot, R., et al., 2021. Adherence to the test, trace, and isolate system in the UK. BMJ 372, 608.

Verwilt, J., Hellemans, J., Sante, T., Mestdagh, P., Vandesompele, J., 2022. Evaluation of efficiency and sensitivity of 1D and 2D sample pooling strategies for SARS-CoV-2 RT-qPCR screening purposes. Scientific Reports 12, 6603. 10.1038/s41598-022-10581-6

World Health Organization, 2026. CORC-CoV Coronavirus Research & Development Roadmap.

